# “The expediency of local modelling to aid national responses to SARS-CoV-2.”

**DOI:** 10.1101/2020.05.27.20107656

**Authors:** Bernard M. Groen, Paul Turner, Peter Lacey

## Abstract

**Background:** With the SARS-CoV-2 pandemic gripping most of the globe, healthcare and economic recovery strategies are being explored currently as a matter of urgency. The underpinning rationale of this paper is that we believe that health and care services are provided locally, therefore, local implications of national policy need to be reflected when informing national responses to the SARS-CoV-2 pandemic.

**Methods:** We adopted the assumptions underlying the United Kingdom government’s national epidemiological model which influences the national policy response to the SARS-CoV-2 pandemic. We used these in a local context and show projections in terms of presentations of symptomatic patients differ in a variety of settings. **Setting:** *North of England, United Kingdom, population modelled at four local constituent levels which aggregated gives a total population of 3.2m*.

**Results:** We clearly demonstrate that there is significant difference in the way the national modelling outputs are replicated at local levels. Specifically, in terms of projected increased levels of demand for services on the local health and care systems.

**Conclusions:** We present significant evidence of differing timelines specifically in terms of subsequent projected peak demands. Additionally, it clearly indicates varying levels of such demand throughout the four modelled localities. These idiosyncrasies are ‘masked’ by both regional and national approaches to modelling. We urge readers to ensure that any national policy is appropriately adopted through the use of complementary bottom up approach, to suit local health and care systems. Finally, we share our methodology to ensure other professionals could replicate this study elsewhere.

## Background

National governments are forced to take urgent action using national policies to restrict social interactions and temporary business closures which impact significantly on numerous factors of our lives. In order to fully understand how limited health and care resources could be utilised and maximised in response to this pandemic, we need to create a thorough understanding of the impact of national policy on local health and care systems. Our guiding principle and understanding here is that health and care services are provided locally (effectively at Integrated Care Partnership – ICP level^4^) not regionally or nationally, indeed, not in aggregate but to individuals. Therefore, we conducted a study using well-established model parameters which are used to inform national policy and adapted these to consider local nuances to see whether any projected model outputs would behave differently to national projections. We show how projections in terms of presentations of symptomatic patients in a hospital setting significantly vary between local communities. Indeed, when aggregated up to regional or national levels, such local idiosyncrasies fade which may have profound consequences in efforts to coordinating local health and care services in response to SARS-CoV-19.

## Methods

### Model design

A System Dynamic Model (SDM) approach is selected and a Susceptible, Exposed, Infectious, Recovered, (SEIR) stock and flow model designed by the SDM software manufacturer^5^ is adapted for the base model. The model is run over a one-year period starting from 1^st^ January 2020 and calculates one transaction per day using the Euler integration method. SDM offers communities a tool with which to understand their systems and become ready to influence and engage with real-world actions (Minyard et al., 2018) whilst avoiding discrete operational level interference. The purpose of the SDM allows Integrated Care System (ICS) communities to simulate scenarios for non-pharmaceutical interventions to SARS-CoV-2 and examine care provision away from a national perspective. Porter and Oleson discuss limitations in SEIR arising from exponential distribution of latent and infectious times (Porter & Oleson, 2013) and SDM was selected to allow the dynamic application of stochastic distribution of infection of the susceptible population over time. SDM also allows the ability to apply variables to control the transmission of the virus (social distancing measures) with which to test interventions (Lopez & Rodo, 2020) (Milne & Xie, 2020).

The model concept is shown in figure 1 below.

**Figure 1:**
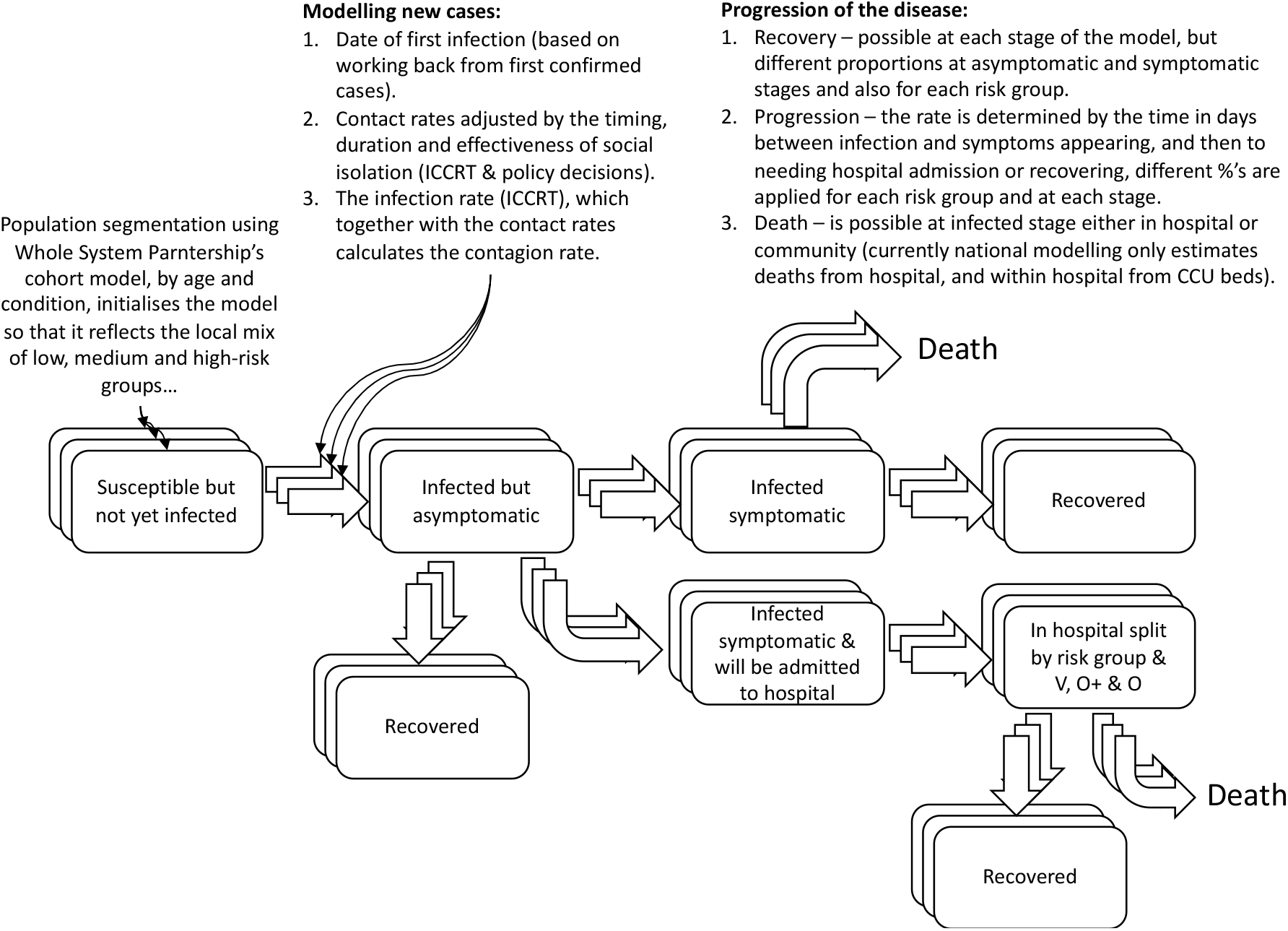
epidemiological model concept

### Data

The SDM is calibrated with data describing the population, virus characteristics and actual outcomes. Population data are extracted from the Office for National Statistics (ONS) census projections by age and Clinical Commissioning Group (CCG) and applied to the ICS or population being modelled (see Localisation sector for further work). The virus characteristics are taken from the Imperial College COVID-19 Response Team (ICCRT) simulation, which informs United Kingdom government’s national policy on the SARS-CoV-2 pandemic (Ferguson et al., 2020); namely the fatality and virus transmission rates by age group. Actual outcomes, for daily hospital deaths and admissions are taken from NHS England daily reports which are consistent with reported hospital length of stay data are extracted from each acute hospital trust. Generic processed data is available by consulting (Groen & Turner, 2020).

### Validation

To maintain fidelity to the ICCRT simulation, the SDM was initially calibrated with United Kingdom population data and the outputs from both models matched (R_o_ = 2.4, the estimated reproduction number at the time of validation). Both models temporally match for expected deaths and critical care bed provision based on proposed intervention efficacy.

### Adaptation and Localisation

Using the ICCRT simulation, the SDM categorised the initial susceptible population into age groupings. To reflect local characteristics, this was augmented to include a risk profile (low, moderate, and high) using population-based health profiles taken from a separate proprietary model (the Cohort model). The cohort model uses data extracted from ONS and the Kent Integrated Dataset to create prevalence and incidence for morbidities (including expected multi-morbidities) adjusted for each English CCG. Models were calibrated for each ICS in the North East and North Cumbria region of England, based on its constituent CCGs, and was seeded for initial infections 30 days prior to the first cluster of deaths on sequential days and validated so that modelled outcomes for ICS hospital bed occupation and deaths fitted actual situation report results. An example ICS results are shown in figure 2 and 3.

**Figure 2:**
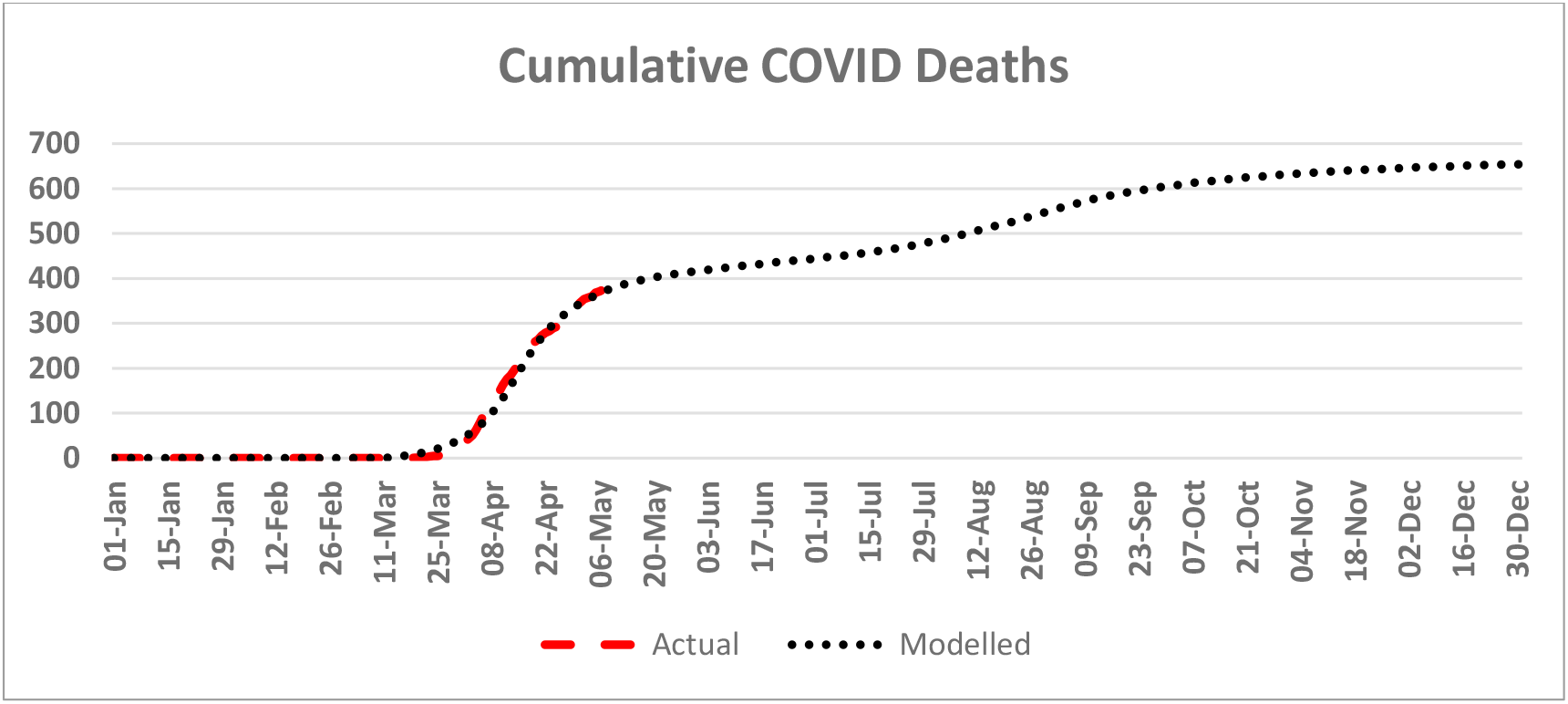
Model validation example 1

**Figure 3:**
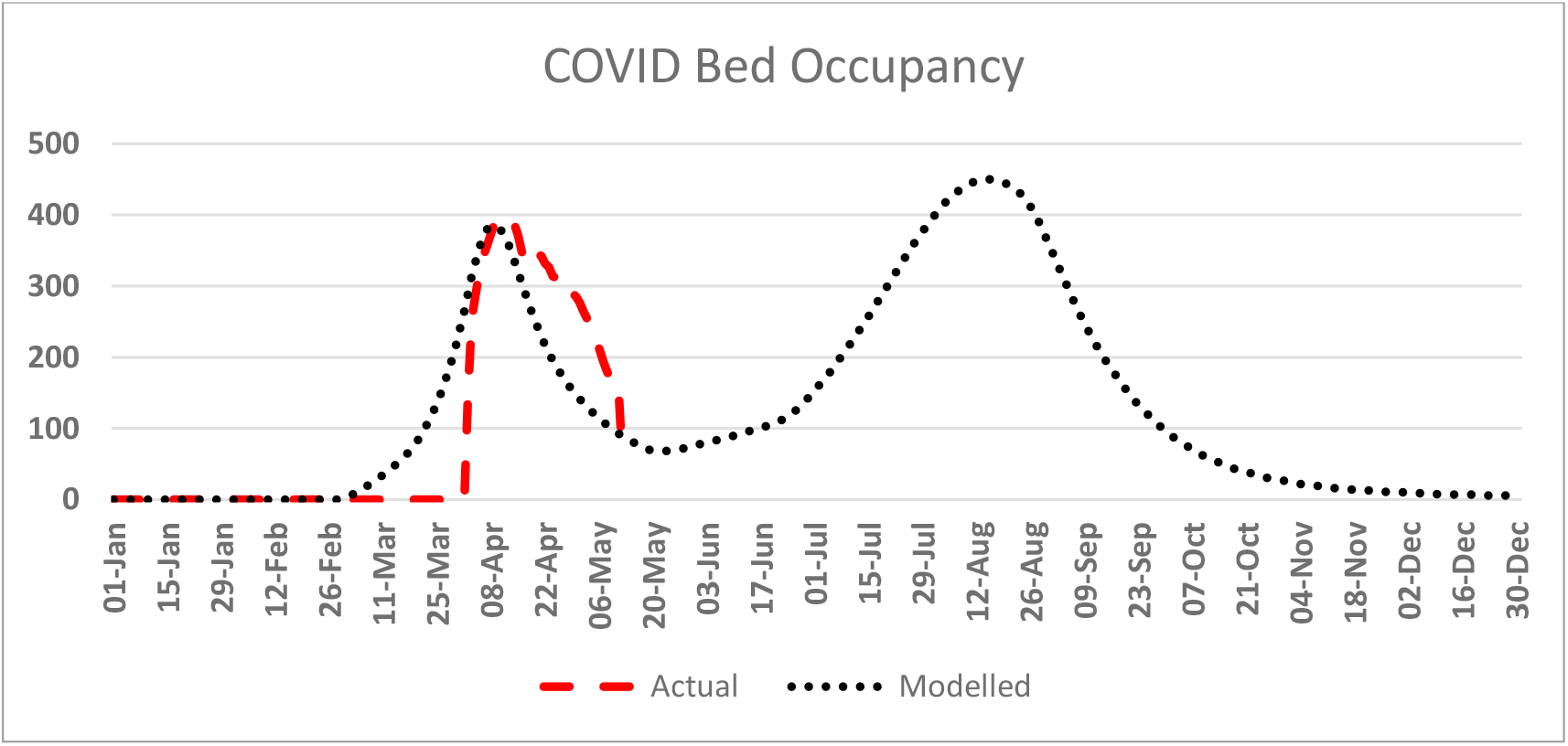
Model validation example 2

### Scenarios

We set four scenarios to simulate the demand on healthcare service provision following the initial intervention to reduce potential virus transmitting contacts introduced by the UK government during March 2020. Baseline contacts pre-intervention (before March 2020) were set at a nominal value of 100 and adjusted for likely reductions from government interventions for the remainder of 2020. The scenarios tested. The adjusted contact value and timings for these scenarios are summarised in table 1 below.

**Table 1:** Summary of scenarios modelled.

| Description |  |
| --- | --- |
| Scenario 1 | The initial impact from the intervention is extended but weakens following initial success. Later in the year the impact from the intervention increases as a result of effective test, track, and trace. |
| Scenario 2 | Gradual, medium to long term relaxation of social distancing from May 10th results in a reduction in the effectiveness of social distancing. |
| Scenario 3 | Cyclical relax/renew - May 10th relaxation with a reduction in the effectiveness of social distancing following by subsequent lock down and release. |
| Scenario 4 | Extended social distancing but weakening of the initial lock down followed by effective implementation of track and trace. |

**Table 2:** Model scenarios for contact adjustments during 2020.

|  | Risk Profile | Jan | Feb | Mar | Apr | May | Jun | Jul | Aug | Sep | Oct | Nov | Dec |
| --- | --- | --- | --- | --- | --- | --- | --- | --- | --- | --- | --- | --- | --- |
| Scenario 1 | Low | 100 | 100 | 80 | 80 | 65 | 65 | 55 | 65 | 65 | 65 | 65 | 65 |
|  | Medium | 100 | 100 | 85 | 85 | 75 | 75 | 65 | 75 | 75 | 75 | 75 | 75 |
|  | High | 100 | 100 | 90 | 90 | 80 | 80 | 75 | 85 | 85 | 85 | 85 | 85 |
| Scenario 2 | Low | 100 | 100 | 80 | 80 | 60 | 60 | 60 | 60 | 60 | 60 | 60 | 60 |
|  | Medium | 100 | 100 | 85 | 85 | 70 | 70 | 70 | 70 | 70 | 70 | 70 | 70 |
|  | High | 100 | 100 | 90 | 90 | 75 | 75 | 75 | 75 | 75 | 75 | 75 | 75 |
| Scenario 3 | Low | 100 | 100 | 80 | 80 | 50 | 50 | 60 | 50 | 50 | 60 | 60 | 60 |
|  | Medium | 100 | 100 | 85 | 85 | 60 | 60 | 70 | 60 | 60 | 70 | 70 | 70 |
|  | High | 100 | 100 | 90 | 90 | 75 | 75 | 80 | 75 | 75 | 75 | 75 | 75 |
| Scenario 4 | Low | 100 | 100 | 80 | 80 | 65 | 65 | 70 | 70 | 70 | 70 | 70 | 70 |
|  | Medium | 100 | 100 | 85 | 85 | 75 | 75 | 80 | 80 | 80 | 80 | 80 | 80 |
|  | High | 100 | 100 | 90 | 90 | 80 | 80 | 80 | 80 | 80 | 80 | 80 | 80 |

### Results

The scenarios were run for four local communities in the North East of England and North Cumbria. All calibration data were assumed the same except for population health needs and the seeding date required for timing in order to fit the model outcomes to actual. Modelled Outcomes from the scenarios were analysed for hospital deaths, acute hospital bed occupancy split between bed capacity requiring ventilation, continuous positive airway pressure (oxygen plus) and oxygen. Epidemiological progression was also analysed using:

- the effective reproduction number - R_e_ (the proportion of overall population remaining susceptible to the virus). R_e_ = R_o_ x (population susceptible / total population)
- the effective reproduction number over time - R_t_ (representing the average new infections arising from active infections at time).

### Epidemiology

The differences in susceptible population resulting from the pre-intervention viral spread can be seen in the results in figure 4. The efficacy of the changing contact rate arising from the scenarios creates dynamics in viral progression, for example West (i.e. North Cumbria) recorded earlier cases in larger quantities than the other areas within the ICS and the effect can be seen in R_e_ lowering at a more rapid rate in the period to April 2020.

**Figure 4:**
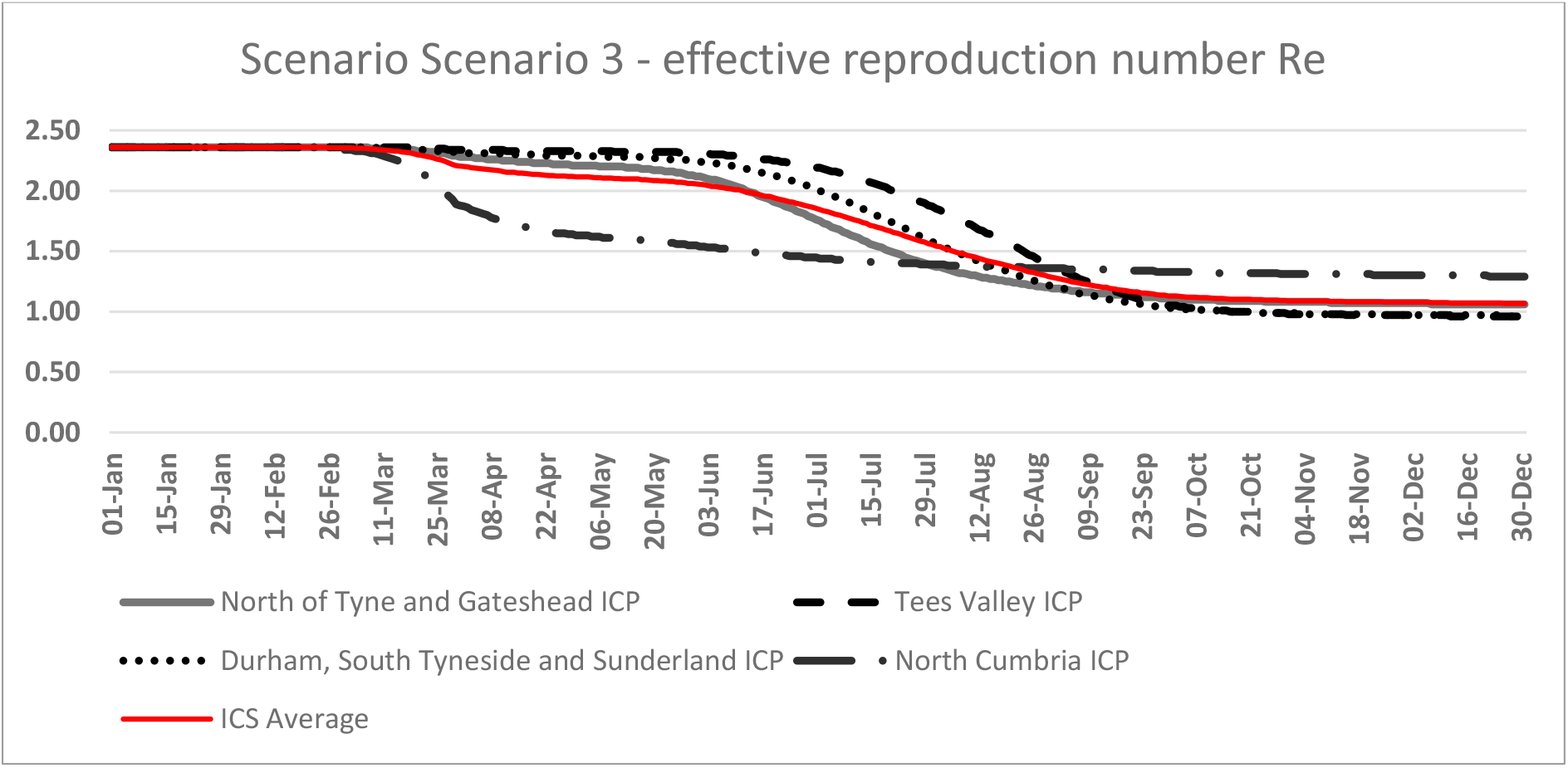
Differences in effective reproduction number R_e_ under scenario 3

**Figure 5:**
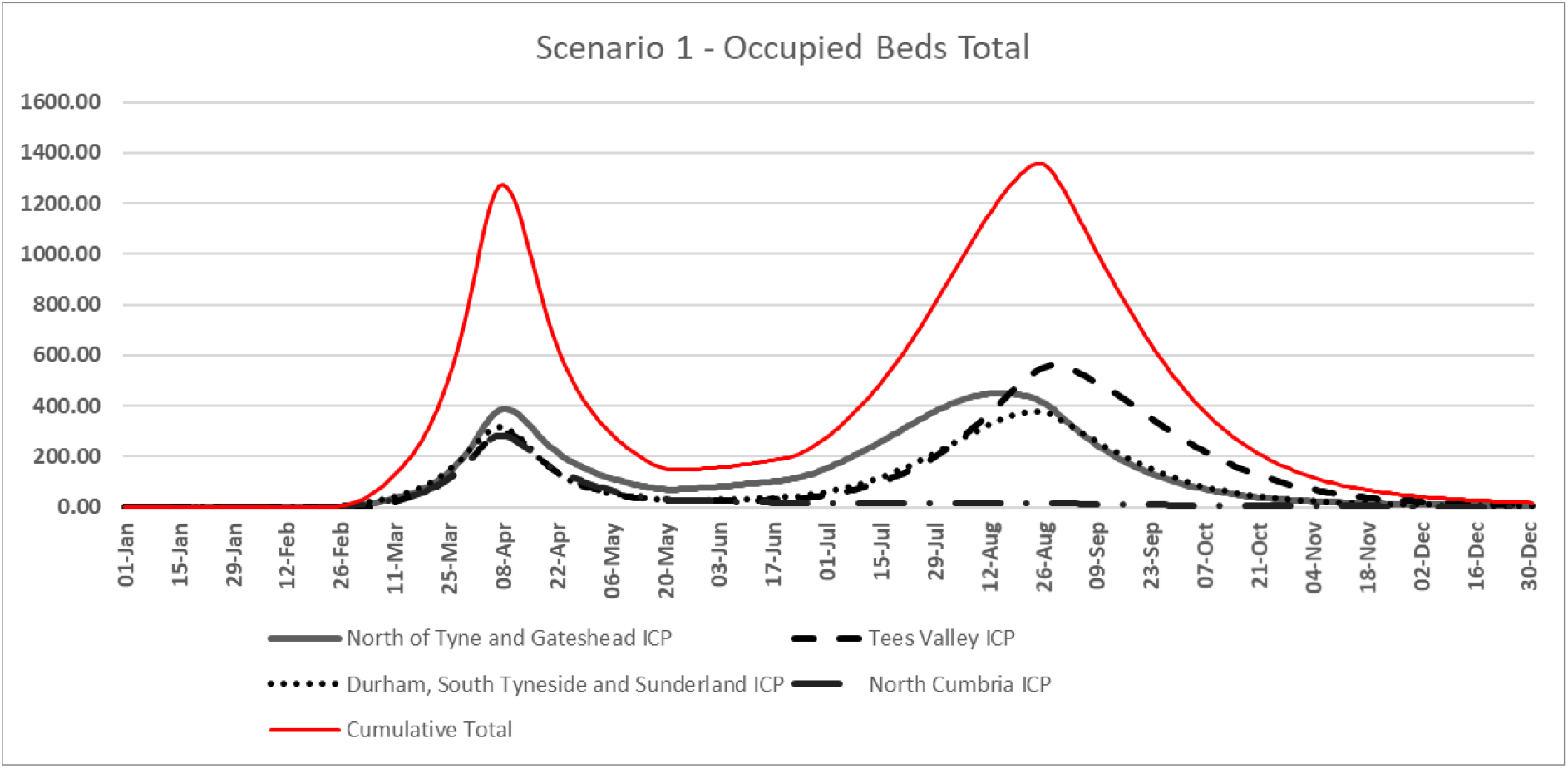
Showing scenario 1 results for the entire ICS then split by ICPs clearly showing variation between local communities.

The dynamic of viral progression can be seen in the demand for acute hospital beds and deaths in the following sections, modelled outputs for all four scenarios are fitted to actual performance from the model inception until the data of simulation in mid-May 2020.

Results above are split by locality in figure 6 modelling demand for oxygen beds. This illustrates overall demand for this type of bed, however, we have modelled this for each type of SARS-CoV-19 demand a more detailed analysis and anonymised data can be accessed at (Groen & Turner, 2020). Generally speaking, the demand of beds is based on the infection rate amongst the constituent population remaining susceptible to the virus, which accounts for the higher number of total demand for such bed types. However, it is important to note that patients admitted to this type of beds tend to have a shorter length of stay and tend to be clinically less complicated to manage operationally.

**Figure 6:**
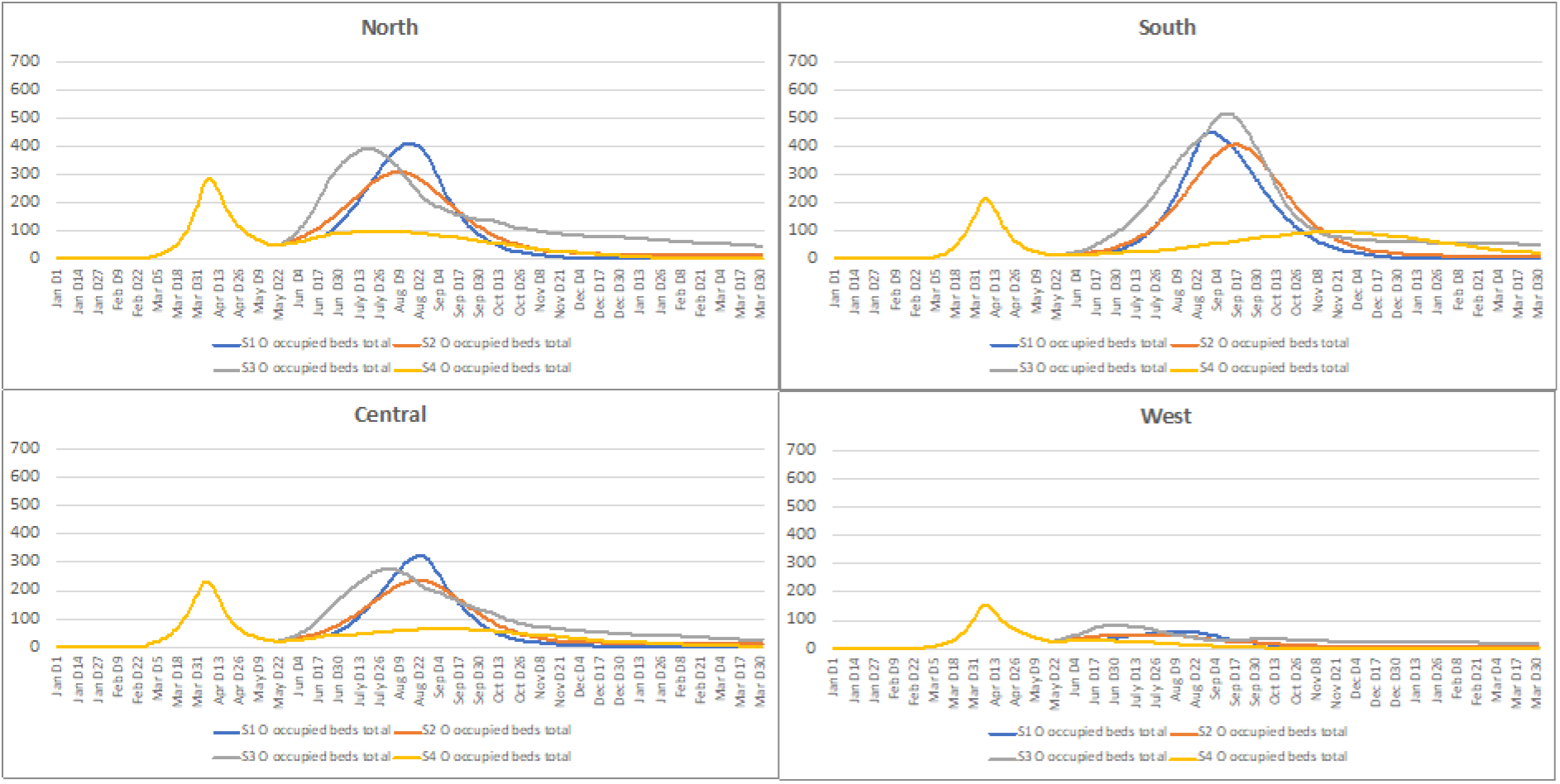
Showing scenario 1 results demand for oxygen beds by individual ICP, clearly showing local idiosyncrasies in terms of demand.

**Figure 7:**
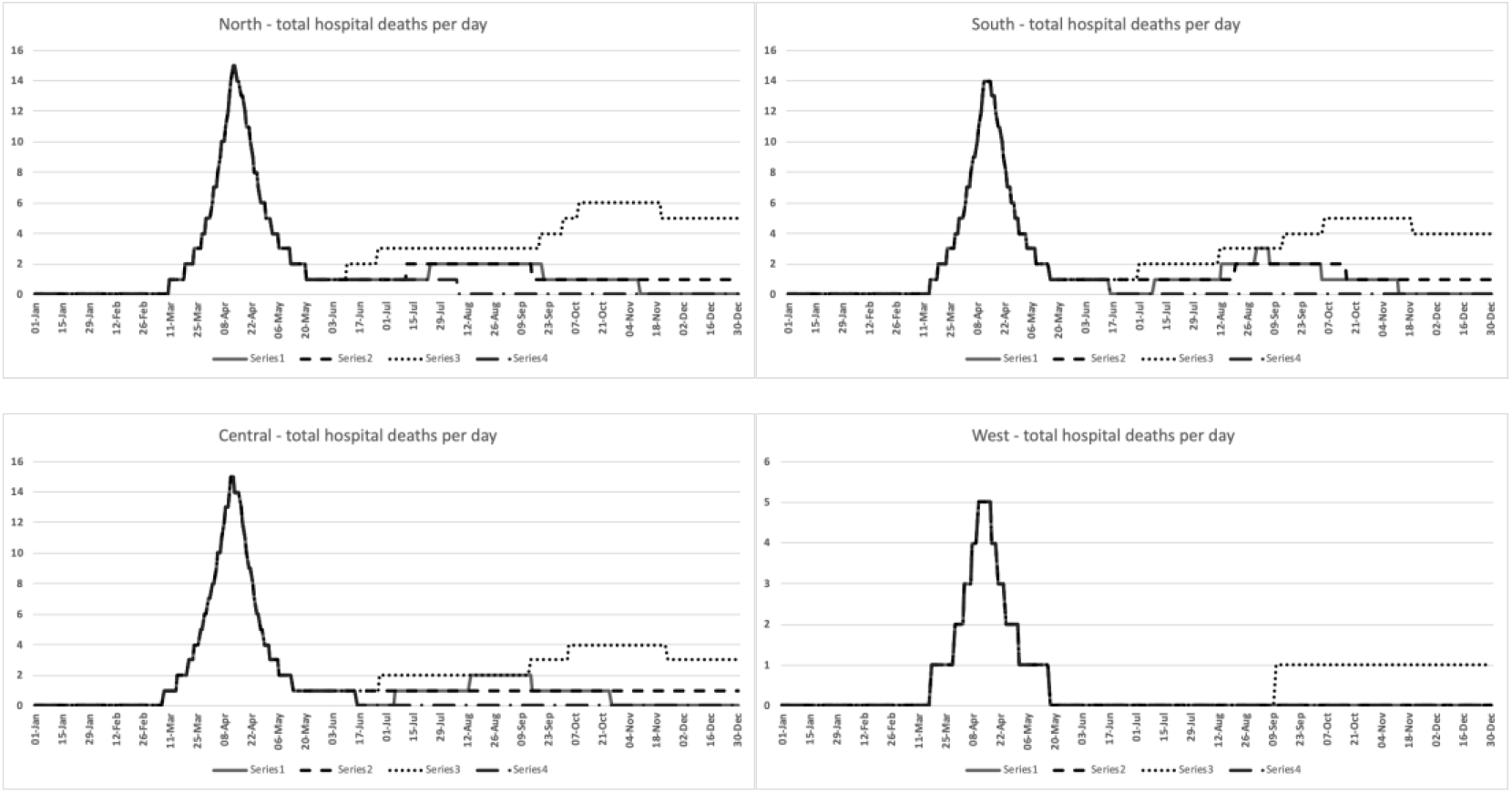
Deaths in hospitals modelled

The model is indicating a higher first peak in terms of hospital deaths, owing to the fact that the cohort approach considers the remaining susceptibility amongst the uninfected population and stratifies this by category of cohort.

## Discussion

With regards to contact rate and seeding of the model – we have subjected our approach to numerous tests to demonstrate that the overall outcomes from the pandemic, for example in terms of deaths, are highly sensitive to the timing of initial seeding and the subsequent lock-down. We applied the national model on a single data file on the 23rd March 2020. In areas of high initial seeding of the virus the additional days of spread may result in differences in the overall number of deaths expected and in the subsequent size of any second wave of the pandemic. This will rely on different levels of the remaining susceptible populations after the observed initial peak and potential projected ones, see (Biskup & Prewitt, 2020) for a further discussion. We certainly believe that this strengthens the case for scenario planning of subsequent easing or resumption of any lock-down measures on the basis our locally focused approach to modelling this pandemic.

The model is sensitive to two major variables outside of the virus characteristic, these are seeding volume/date and the contact reduction. Model seeding assumes a certain number of infections entering the region to create the virus uptake prior to the first peak, however it does not currently assume any new cases coming from out of region after that time. This seems an unlikely scenario and future version of the model will address this issue as more evidence on travel following easing of government policy is made we aim to refine this using intelligence and conceptual approaches such as outlined in (China CDC, 2020). This will be important as the current epidemiological element of the model is one of self-perpetuating transmission – the current infected population are the only ones who will contribute to new infections. Social distancing contacts also depend on a less aggregated view. Local characteristics for potentially virus transmitting contacts can be estimated through data including urban / rural or population density measures (Leung et al., 2020; Li et al., 2020), household density, vulnerability indices and movement/transport (Kraemer et al., 2020) see (Prem et al., 2020) for how these factors played a role in China’s approach to social isolation. This would allow greater understanding of influences on clinical demand through more sophisticated scenario building.

As per well-established literature, the model concurs on recent peer-reviewed publications and popular media outlet coverage which draw attention to a ‘second wave’ scenario, see (World Health Organization, 2020) and (Xu & Li, 2020) for example. Indeed, it is within that context that we stress the importance of locally defined modelling approach (effectively; bottom up) which reflects local population needs, which, when aggregated will comprise a more insightful and nuanced approach to inform national approaches to this global challenge. We call on researchers to adopt our approach within their own local context to ensure health and care demands are met locally within the inevitable constraints that comes with national policy.

## Notes on Limitations

Contact too sensitive – developed into multidimensional transmission by place, movement vulnerability, social/domestic contact which will be applied to the age-based population health needs. Questions over the small number needed for seeding compared to local estimates (may mean infection rate is higher as per other papers). Model fitted to recorded hospital admissions and deaths (although COVID deaths outside of hospital are modelled.

- Contact rate and seeding – highly sensitive independent factors
- Deaths peak in first wave – more high risk / vulnerable (from cohort model)
- Beds have generally higher demand in second (subsequent waves, depending on relaxation in scenario)
- Time of initial infections important in epidemiology.
- The capacity numbers used in the reports remain high as they are in use nationally at this level but we will look to work with providers to add more ‘accurate’ numbers in the coming week, alongside those currently in use.

## Data Availability

Further methodological approach details available upon request from lead author. Data is available on Harvard DataVerse, see link below.

https://doi.org/10.7910/DVN/MTK2NW

## Footnotes

4 Note on geography: The ICS covers the North East and North Cumbria of England. There are four (4) constituent ICPs; 1) North (North of Tyne and Gateshead ICP); 2) Central (Durham, South Tyneside and Sunderland ICP); 3) West (North Cumbria ICP); 4) South (Tees Valley ICP). Total population ~3.2m.

5 iSee Systems https://www.iseesystems.com/

## Notes

### Competing Interest Statement

Dr Turner discloses that he consults for NHS partners through WSP, in addition to his role at the University of Lincoln.
Mr Lacey discloses that he consults for a variety of NHS organisations through WSP.

### Funding Statement

This research received no specific grant from any funding agency in the public, commercial or not-for-profit sectors. Time and resources were made available by NHS Health Education England.

### Author Declarations

NHS Health Education England Durham University

